# Effectiveness of Propofol in Mitigation of Emergence Agitation Among Pediatric Patients Undergoing Elective Surgery Under General Anesthesia: Prospective Cohort Study

**DOI:** 10.1101/2025.01.07.25320109

**Authors:** Kuchulo Geremu Gelgelo, Orissa Orkissa Tessema, Wondu Reta Demissie, Gezahegn Tesfaye Mekonin, Kanbiro Gedeno Gelebo

## Abstract

**Introduction:** Emergence agitation is the abnormal mental status that develops as the result of anesthesia administration during the transition from unconsciousness to complete wakefulness. It occurs in all age groups but is more common in pediatrics. If emergence agitation is not prevented, it can lead to serious life-threatening phenomena. Thus, preventing the occurrence of emergence agitation would reduce the unwanted side effects and makes emergence smooth.

**Objective:** This study aimed to assess the effectiveness of propofol in the mitigation of emergence agitation among pediatric patients undergone elective surgery under general anesthesia at Jimma University Medical Center.

**Methods:** A prospective observational cohort study was conducted from October 1/2022 to December 30/2022 among pediatric patients aged 2-14 years who underwent elective surgery under general anesthesia. A total of 76 patients were enrolled in the study in two groups (propofol and non-propofol) with 38 patients in each. Data were collected from all study populations who fulfilled the inclusion criteria. The incidence and severity of EA were assessed by the pediatric anesthesia emergence delirium scale in the post-anesthesia care unit at different time intervals. The presence of emergence agitation was considered when the pediatric anesthesia emergence delirium scale was ≥ 12 and severity was classified as no agitation [0–11], agitation [12–15], severe agitation [≥ 16]. The data were checked manually for completeness then entered and cleaned using Epi Info, version 7, and exported to Statistical Package for Social Sciences (SPSS) software, version 26.0 for analysis. Normality was tested using the shapiro-wilk test, and histogram. A comparison of numerical variables between the study groups was done using an independent sample t-test. Chi-square/Fischer exact test was used to compare categorical variables and presented as numbers and percentages. The incidence of EA was also presented in confidence interval and relative risk. A p-value less than 0.05 was taken as statically significant.

**Result:** The overall incidence of EA in both groups was 48.7% (37%−60.4%). The incidence of EA in the propofol group and non-group was 34.2% (19.6%−51.4%) vs 63.2% (46%−78.2%) respectively p=0.012, which means that EA was significantly decreased in the propofol group in comparison to the non-propofol group. There was a significant reduction in the severity of EA at time intervals of 5th and 15th minutes; P =0.012 vs 0.029 respectively. However, there was no significant difference in the severity of EA at the 30th minute between the groups; P=0.08. The relative risk of EA in the propofol group was 0.54; 95% confidence interval (CI) 0.3270.896. This showed propofol group had less risk of emergence agitation in comparison to the non-propofol group. Conclusion and Recommendation: The administration prophylactic propofol at the end of surgery is vital in reducing both the incidence and severity of emergence agitation and makes recovery smooth. Thus, we recommend the routine prophylactic propofol at the end of surgery to reduce the incidence and severity of postoperative emergence agitation in pediatric patients.

## INTRODUCTION

General anesthesia (GA) is a drug-induced reversible state of unconsciousness, amnesia, analgesia, and immobility. It is maintained by a combination of intravenous agents, inhalational agents, opioids, and muscle relaxants throughout the surgery. Emergence from GA is the stage of GA featuring the patient’s progression from unconsciousness status to wakefulness and restoration of consciousness [1]. Emergence from GA is accompanied by different complications and classified based on the organ affected such as: cardiac, respiratory, renal, and neurological [2]. From neurological complications, emergence agitation (EA) is a well-known phenomenon in pediatric anesthesia and was first described by Eckenoff as abnormal mental states characterized by confusion, irritability, disorientation, inconsolable crying, restlessness, and non-purposeful physical movements [3].

The precise pathophysiological mechanism of EA after GA is unknown [4]. During emergence from GA, thalamo-cortical connectivity in sensory networks, and activated midbrain reticular formation are preserved. However, delayed recovery of impaired functionality of subcortical and thalamo regulatory systems could contribute to the defects in cortical integration of information, which could lead to confusion or an agitated state [5]. In children, the proposed causes of EA include high levels of anxiety regarding surgery, new environments, separation from parents, encounters with unfamiliar medical staff, and immature neurological activity [6].

There are a lot of factors contributing to the development of EA and classified as: Patient related factors such as Age, maturation status, and preoperative anxiety; Surgery related factors such as type of procedure and anesthesia-related factors such as usage of modern inhalational anesthetic agents like sevoflurane, isoflurane, and halothane are common causes of EA [7]. EA can occur in all age groups but is more common in pediatrics with an incidence of around 10%-80% [8]. Children who received volatile anesthetics (sevoflurane and desflurane) as maintenance are at high risk of developing EA. A reported incidence of EA in children maintained with sevoflurane has ranged from 24-66% and increased to 80% in preschool children [9]. Once the EA has occurred it results in a lot of negative consequences like self-injury, removing their catheters, drains, intravenous cannula, falling out of bed, bleeding at the surgical site and prolong recovery time in the post-anesthesia care unit (PACU). It predisposes their parents or caregivers to anxiety, stress, and increasing medical care costs [10]. Several scales have been proposed for assessing EA in children but they were not been tested psychometrically and they follow emotional distress and psychomotor agitation as surrogate markers of delirium. Among them, the pediatrics anesthesia emergency delirium (PAED) scale which has been tested psychometrically is the most sensitive and specific for assessing emergency agitation for children ≥2 years of age [11].

For EA prevention is preferred over treatment, and many prophylactic medications have been used to decrease the incidence of EA including fentanyl, ketamine, midazolam, and most recently dexmedetomidine [12,13]. Propofol which is an intravenous hypnotic agent has also been used as an adjuvant or alone for the prevention of EA [14]. It works by enhancing γ-aminobutyric acid (GABA) activated chloride channels with a blood-brain equilibration half-life of approximately 2 to 3 minutes and a distribution half-life of 2 to 4 minutes. So that due to its rapid onset and the short duration of action, patients can rapidly recover from it. In addition to this, it has also calm, anticonvulsant activity, antipruritic effects, and antiemetic properties [15].

EA is the acute confusion state during recovery from GA and occurs both in the operation room (OR) and PACU. The patients present with disorientation, irritability, confusion, restlessness, thrashing, and crying [16]. EA is a well-documented clinical phenomenon, particularly in the pediatric population, and prevalent in early to mid-childhood with an incidence of (10%-80) [17]. Poorly prevented EA not only impacts the children negatively but also the parents and healthcare providers [18]. The children are harmed both physically and psychologically and mainly includes: accidental removal of drains or dressings, infection, falling out of bed, bleeding at the surgical site, prolong recovery time in the post-anesthesia unit and discharge from the hospital, nightmares, eating problems and increased fear of physicians [19].

On the other hand, EA stimulates the sympathetic system and consequently increases the heart rate and blood pressure and giving rise to complications such as brain and heart damage and also delaying wound healing [20]. Not only children but also caregivers such as family, nurses, and anesthetists are harmed. Especially it causes physical injury as well as a load of giving extra care to healthcare providers and also decreases the satisfaction of the family.

Not only this but also causes financial burdens to the hospital because once the children are agitated they may damage the monitoring and other healthcare facilities and also increase the number of staff required to give care for the average length of time spent with these patients, these all thing leads to additional costs for the hospital [21].

Different measurements have been taken for the prevention of EA and from them, a sub hypnotic dose of prophylactic Propofol has been used. A randomized control trial (RCT) was conducted on the administration of Propofol at the end of Surgery for the prevention of EA and showed that the incidence of EA was significantly lower in the propofol group compared with the saline group (19.5% vs. 47.2%; P = 0.01) respectively [22]. A double-blind study was conducted on assessing the effect of Propofol administration in the prevention of EA at the end of sevoflurane anesthesia for magnetic resonance imaging (MRI) and reported that the incidence of EA was lower in the propofol group and not shown to delay recovery or discharge time [23].

Another double-blind study was conducted on the effectiveness of Propofol in decreasing the incidence and severity of EA in children undergoing adenotonsillectomy and finally concluded that propofol has no effect in reducing the incidence and severity of EA [24]. The reason why propofol is not effective is postulated as because sevoflurane-related EA is affected by a combination of surgical procedures rather than the impact of propofol alone. A comparative study conducted on dexmedetomidine and Propofol in the prevention of postoperative incidence and severity of EA showed that administration of dexmedetomidine was more effective than Propofol 1 mg/kg in decreasing the incidence and severity of EA when given before the end of surgery [25].

To manage agitated child’s, anesthetists use analgesics and sedative drugs. However, those techniques prolong the duration of the patient’s stay in the PACU and not only also miss diagnose EA as Pain, and administering unnecessary medications to the child increases cost and drug exposure [26–29]. Even though some literatures support the effectiveness of Propofol for the prevention of EA, others aren’t. Most of studies were done on specific surgical type and none of the kinds of literature were conclusive on the effectiveness of propofol in the prevention of EA in respective of different surgical procedures [30]. So, we intended to conduct this study by including all surgical procedures that have been done in pediatric patients in our setup to show the effectiveness of propofol in the mitigation of EA in respective of different surgical procedures.

Evidences from different literatures show agitated child removes drains or dressings accidentally, infection, falling out of bed, bleeding at the surgical site and can cause damage to equipment such as monitoring (blood pressure apparatus, pulse oximetry, electrocardiograph, and bed). Besides, agitated patient increases stress and workload on anesthetist, PACU nurses and other care givers by precluding them not to provide care to other patients simultaneously. Furthermore, it increases stress, and financial expense and decrease satisfaction of family or guardian. We hypothesized there is no difference in occurrence of postoperative emergence agitation between two groups. Hence, the purpose of this study is to assess the effectiveness of propofol in the mitigation of EA among pediatric patients undergoing elective surgery under GA.

## MATERIALS AND METHODS

Jimma University, institute of health, faculty of medical sciences, institute review board granted ethical approval for this study on September 29, 2022 with protocol number JUIH/IRB/81/22, Jimma, Ethiopia. Family/legal guardian of participants provided written informed consent.

The study was registered in the Research Registry with unique identity number: researchregistry11034 and linked at https://www.researchregistry.com/browse-the-registry#home/.

This study has been reported in line with the STROCSS criteria [31].

We conducted institutional-based prospective cohort study among pediatric patients underwent elective surgery under GA from October 1/2022 to December 30/2022. All elective ASA I & II and age of 2–14 years old pediatric patients underwent elective surgery under general anesthesia were included. Patients with a known psychiatric disorder were excluded.

The Sample size was estimated from the previous study done in Addis Ababa, Ethiopia [32]. According to their study, the incidence of emergency agitation in propofol and non-propofol group was 31.1% and 64.4% respectively. The Epi info version 7 stat calculator was used for sample size calculation. It was calculated by assuming the ratio of unexposed to exposed 1:1, study power of 80% and confidence interval of 95%. The calculated sample size for propofol and non-propofol group was 38 patients for each.

Data were collected from all patients who fulfilled the inclusion criteria during study period. After a brief explanation about the study for their parents or guardian, informed written consent was obtained from parents or guardian, and only those who agreed were included in this study.

The trained data collector has been in OR and documented the type of medication used for both induction and maintenance by the anesthesia providers. To make the recovery from anesthesia smooth some anesthesia providers used propofol as prophylaxis at the end of surgery and some did not. So, the data collectors strictly observed and documented it.

After the patient was transferred to the PACU the presence and severity of EA was assessed and documented by trained data collector by using the PAED scale at time interval of 5th, 15th, and 30^th^ minutes. This scale is the most sensitive and specific for assessing EA and it consists of 5 criteria that are scored using a 5-point scale, then the results of each item are converted into scores and summed up out of 20.

During data collection, the data collectors filled each parameter at the proposed time interval and summed the total score. The investigators collected the filled questionnaires and classified the patients based on the above stratification. If the child was asleep at the time of measurement, it was considered as “not agitated.”

The presence of EA was defined when the PAEDS ≥12 and the severity was stratified as 0-11 no agitation; 12-15 agitation and score ≥16 severe agitations [33,34].

ASA status: is surgical risk stratification validated by the American Society of Anesthesiologists physical status; described as follows:

ASA I: a healthy patient with no organic/physiological/ psychotic problems.

ASA II: patients with controlled medical conditions with mild systemic effect and no limitation of functional ability.

EA: Its presence was defined when the PAEDS score is ≥12 and for the severity when the score is ≥ 16.

The data were checked manually for completeness and the entered and cleaned using Epi Info, version 7, and exported to Statistical Package for Social Sciences (SPSS) software, version 26.0 for analysis. The normality of the data was checked by the Shapiro-Wilks test and histogram.

Independent student t-test was used to compare continuous variables between study groups and was reported as mean ± standard deviation. Chi-square/Fischer exact test was used to compare categorical variables. The incidence of EA was presented in number (percentage), confidence interval (CI), and relative risk (RR). A p-value less than 0.05 was taken as statistically significant.

## RESULTS

### Sociodemographic and preoperative profiles of the respondents

A total of 76 pediatric patients between 2-14 years old who underwent surgical procedures under GA were enrolled in the study. All patients who were initially enrolled completed the study. The patients enrolled for the study were divided into two groups’ propofol and non-propofol with 38 patients in each. There was no significant difference in the descriptive analysis of the patients found, with the distributions of age, sex, weight, and physical state (ASA class) found (Table 1). The distribution of population in each group was male 23(60.5%) and female 15(39.5%) in the propofol group and male 20(52.6%) and female 18(47.4%) in the non-propofol group.

**Table 1.** Sociodemographic and preoperative Profiles of the respondents.

| **Variables** | **Description** | **Propofol group** | **Non-Propofol group** | **P-Value** |
| --- | --- | --- | --- | --- |
| **Age in a year** | Mean ± SD | 6.05 ± 3.55 | 7.11 ± 4.2 | 0.2 |
|  | Range | 2-14 | 2-14 |  |
| **Sex; number (percentage)** | Male | 23(60.5%) | 20(52.6%) | 0.6 |
|  | Female | 15(39.5%) | 18(47.4%) |  |
| **Weight in kg;** | Mean ± SD | 17.97 ± 6.6 | 20.1 ± 7.6 | 0.2 |
|  | Range | 10-34 | 10-33 |  |
| **ASA statues;** | I | 38(100.0%) | 30(78.9%) | 0.5 |
|  | II | 0(0.0%) | 8(21.1%) |  |
| **premedication in Number(percentage)** | Yes | 33(86.8%) | 31(40.8) | 0.3 |
|  | No | 5(13.2%) | 7(9.2%) |  |
| **Type of surgery in number (percentage)** | ENT | 6(15.8%) | 2(5.3%) | 0.5 |
|  | Ophthalmic | 10(26.3) | 11(28.9%) |  |
|  | Abdominal | 10(26.3%) | 8(21.1%) |  |
|  | Urology | 4(10.5%) | 10(26.3%) |  |
|  | Orthopedics | 6(15.8%) | 7(18.4%) |  |
|  | Thoracic | 0(0.0%) | 4(10.3%) |  |
|  | Neuro |  | 2(5.1%) |  |

The mean and standard deviation (SD) of age in the propofol and non-propofol groups was (6.05 ± 3.55 vs 7.11 ± 4.2) respectively. The mean and standard SD of weight (Kg) in the propofol group and none propofol group was (17.97 ± 6.6 vs 20.1 ± 7.6) respectively. All the patients who took propofol were ASAI and in both group majority of number took premedication. Ophthalmic surgery was dominated in both groups, 26.30% (10/38) vs 28.90 % (11/38) in the propofol and non-propofol group respectively. Whereas neuro and thoracic surgery were rarely performed in both groups and accounted for the least percentage.

### 2. Intraoperatively used anesthetic agents for the respondents

Analgesics agents were given for all groups and opioids were the dominant analgesic agents used in both groups 65.8% (25/38) vs 72% (28/38) in the propofol group and non-propofol group respectively, Ketofol was frequently used as the induction of anesthesia 36.8% (16/38) in propofol group whereas ketamine in non-propofol group 36.8% (16/38)

In both groups, isoflurane was the most highly used for the maintenance of anesthesia, 73.7 % (28/38) vs 65.8 % (25/38) in the propofol and non-propofol group respectively. The duration of surgery and anesthesia was also presented in mean ± SD. There were no statistically significant differences between the groups as shown in the following table 2.

**Table 2:** Intraoperatively used anesthetic agents, duration of surgery and anesthesia of the respondents.

| Intraoperative agents and duration | Type | Propofol group | Non-propofol group | p-value |
| --- | --- | --- | --- | --- |
| Analgesics | Opioid | 25(65.8%) | 28(72%) | 0.23 |
|  | Non opioid | 9(23.7%) | 4(10%) |  |
|  | Both | 4(10.5%) | 7(18%) |  |
| Inductions | Propofol | 14(36.8%) | 10(26.3%) | 0.8 |
|  | Ketamine | 7(18.4%) | 14(36.8%) |  |
|  | Ketofol | 16(42.1%) | 13(34.2%) |  |
|  | Inhalational | 1(2.6%) | 1(2.6%) |  |
| Maintenances | Halothane | 10(26.3%) | 13(34.2%) | 0.4 |
|  | Isoflurane | 28(73.7%) | 25(65.8%) |  |
| Duration of surgery | Mean ± SD | 69.6 ± 28.6 | 82.24 ± 33.4 | 0.08 |
| Duration of anesthesia | Mean ± SD | 74.32 ± 28 | 87.42 ± 33.9 | 0.07 |
| Values are number (percentage), mean ± SD. No statistical significance between the two groups | | | | |

### 3. Status of EA in both groups and each group

The incidence of EA in both groups was: agitated 48.7% (37−60.4%) and non-agitated 51.3% (39.6−63%). The incidence of EA in each group was: in the propofol group the agitated and non-agitated patients were: 34.2 % (19.6-51.4%) vs 65.8% (48.6%−80.4%); respectively; P=0.074 and in the non-propofol group the agitated and non-agitated patients were: 63.2 % (46−78.2%) vs 36.8% (21.8-54%); respectively; P=0.144, as shown in the figure below.

**Figure 1:**
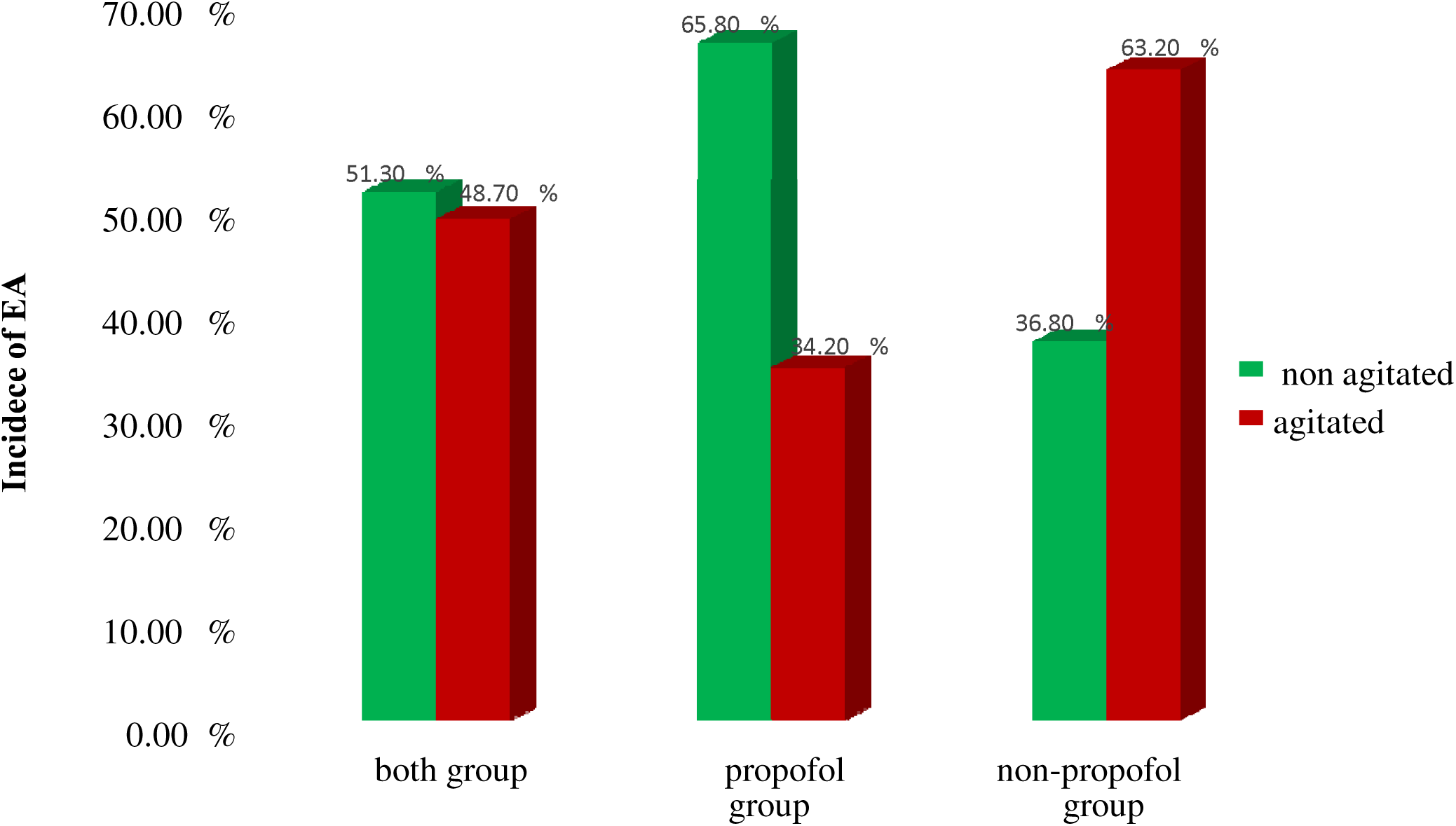
incidence of EA in both and each studied group.

#### A. Severity of EA in the propofol group and non-propofol groups at different time intervals (5^th^, 15^th^ and 30^th^ minutes)

**At the 5^th^ minute**; the severity of EA in the propofol group was: not agitated 28 (73.3%), agitated 8(21%) severely agitated 2(5.3%) and in the non-propofol group was: not agitated 20(52.6%) agitated 10(26.3%) severely agitated 8(21%)); P value =0.011.

**At the 15^th^ minute**; the severity of EA in the propofol group was: not agitated 28(86.8%); agitated 5(13.2%); severely agitated 0(0.0%) and in the non-propofol group was: not agitated 23(60.5%); agitated 14(36.8%); severely agitated 1(2.6%); p=0.029

**At the 30^th^ minute;** the severity of EA in the propofol group was: not agitated 36(94.7%), agitated 2(5.3%) severely agitated 0(0.0%). In non-propofol group was: not agitated 32(81.6%), agitated 6(18.4 %), severely agitated 0(0.0%) with p=0.08.

**Table 3:**
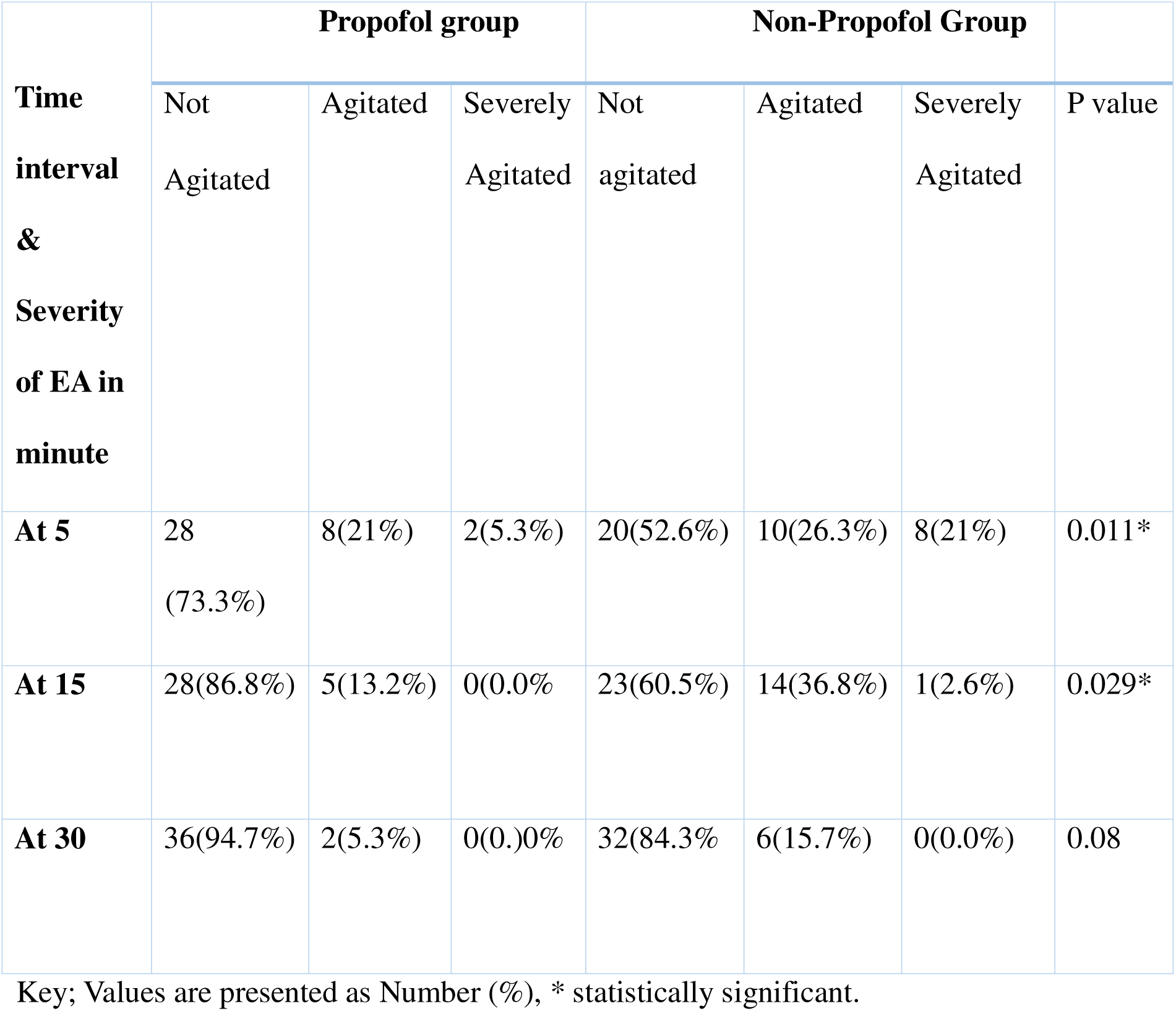
Severity of EA at different time intervals ( 5^th^, 15^th^ and 30^th^ minutes) in the propofol group vs non-propofol group.

**Table 4:** Compare the risk of EA between propofol and non-propofol groups.

| Groups | RR | 95% CI |  | p-value |
| --- | --- | --- | --- | --- |
|  |  | Lower | Upper |  |
| **Propofol group** | 0.542 | 0.327 | 0.896 | 0.012* |
| **Non propofol group** | 1.786 | 1.110 | 2.872 |  |

Key; value is presented as RR (relative risk), CI (confidence interval) and \* statistically significant.

#### B. Effectiveness of prophylactic Propofol administration in reducing the risk of EA

The relative risk (RR) of EA in the propofol group in comparison to the non-propofol group was 0.54; 95% confidence interval (CI) 0.327-0.896, p= 0.012. This means that pediatric patients who took prophylactic propofol had 46% less risk of having EA as compared to pediatric patients who did not take prophylactic propofol at the end of surgery or the risk of EA among pediatric patients took propofol was 0.54 times as high as the risk of EA among pediatric patients did not take propofol.

The relative risk (RR) of EA in non-propofol in comparison to the propofol group was: 1.786; 95% confidence interval (CI) 1.11-2.872); p= 0.012. which means pediatric patients who didn’t take prophylactic propofol had a 78.6% increase in the risk of developing EA as compared to pediatric patients who took prophylactic propofol at the end of surgery or the risk of EA in the non-propofol group 1.78 times as high as the risk of EA compared to the subject who took propofol.

## DISCUSSION

Emergence Agitation is a serious problem in the postoperative period of pediatric anesthesia that can have potential risks to patient safety [10]. This study aimed to assess the effectiveness of propofol in reducing EA in pediatric patients who underwent surgery under General Anesthesia.

In this study, the sociodemographic data of the patients, and preoperative and intraoperative profiles were compared for their proportion among the propofol and non-propofol group and there was no statistically significant difference (p <0.05). This means that the influence of the confounder that might affect the outcome was equivalent in both groups.

In this study, the incidence of EA in the propofol group was 34.2 % with a CI of (19.6%-51.4%). A similar finding was reported by Kim et al. in 2011 in which that the incidence of EA in the propofol group was 48.4% [34]. A comparable Study conducted by Jalili et a. in 2019 showed that the incidence of EA in the propofol group was 41% [33]. Prior research done in 2019 by S. J. Lee et al. reported that the incidence of EA was found to be 50.8% [26] was also consistent with the finding of the current study.

In the present study, the incidence of EA in the non-propofol group was 63.2% with a CI of (46−78.2%). Study conducted by Voepel-Lewis et al in 2003 showed that the incidence of EA in the non-propofol group was 43.2% [16] was also consistent with current findings. This finding was consistent with the present study. Previous conducted in 2008 by Saringcarinkul [8] was comparable to the current study where the incidence of EA was 43.2% in PACU. A study done by Eshetie et al. in 2020 [7] was also consistent with our finding in which the reported incidence of EA was 52%.

In this study, we found that the incidence of EA is higher in the 5^th^ and 15^th^ minute in both groups however none propofol group was more severely agitated than the propofol group. The possible explanation may be the first two-time interval minutes are full of distressing due to the residual effect of anesthesia and facing a new environment. At 30 minute there was no statistically significant difference between the groups in having severe agitation and the possible explanation may be EA is a temporary and self-limiting condition and might not persist till 30 minutes. So, it is important to carefully follow the patient within the first 15 minutes to handle any adverse effects of EA.

In the present study propofol group had less incidence of EA in comparison to the non-propofol group and showed that the incidence of EA in those who took propofol was lowered in comparison with the non-propofol group.

A comparable study conducted in American University of Beirut in 2007 on Strabismus correction showed that the incidence of emergence agitation in the propofol group and the non-propofol group was (19.5% vs 47.2% respectively with a p-value of 0.01 [22]. This may be because we used the same scale for assessing the incidence of EA. However, Aoud et al. assessed both the incidence and severity of EA with a total cut point of 10 whereas we used 12 as the cut point which is the most sensitive and specific for determining the presence of EA.

A similar finding was reported in the previous study conducted by Abushawan et al. on propofol administration at the end of surgery done in children undergoing magnetic resonance imaging (MRI). The incidence of agitation in the propofol group and the non-propofol group was (4.8% vs 26.8%) with a p-value of <0.05 [23]. In comparison to our study, the incidence of EA in both groups is low numerically and the possible reason maybe they used a high cut point (≥ 16) for the presence of EA.

In contrast, a study done in Korea in 2010 on the administration of propofol at the end of the surgery for prevention of emergence agitation, then assessed the statues of EA at T5, T15, and T30 were 61.4%, 27.3%, and 4.5% in the propofol group and in the saline group was 68.2%, 29.5%, and 9.1% respectively and reported that there was no difference in incidence and severity of EA between propofol the groups [24]. Jin Lee et al. case included only adenotonsillectomy surgery and also those maintained with sevoflurane anesthesia. The researcher used Aono’s four-point scale for assessment, which is less sensitive. However, we used PAEDS which is the most sensitive and specific.

RCT study conducted in Indonesia in 2020 reported the incidence of EA was 25.9% in the Propofol group and 51.9% in control group (P=0.006) [35]. Their finding is lower than our finding in which the incidence of EA in the propofol group and non-group was 34.2% and 63.2% respectively (P=0.012). This may be due to difference in study design and their study subjects were those maintained with sevoflurane anesthesia only. In addition, they used Aono’s four-point scale for assessing the incidence which is less sensitive than PAEDS.

A consistent study was carried out in Iran in 2019 on a comparison between the administration of midazolam and propofol at the end of anesthesia for the Prevention of EA. The incidence of EA in the propofol group and saline group was 60% vs 97.6% respectively with p=0.006 [30].

The author concludes that giving propofol near the end of surgical operations may be effective in reducing the severity and incidence of EA in a pediatric patient undergoing tonsillectomy after anesthesia by isoflurane. However, the author did not include all the surgical procedures whereas we included all types of surgery.

A previous study conducted in Penerbit Universiti Sains Malaysia in 2021 compared propofol, ketamine, and midazolam on the incidence and severity of EA. The incidence and severity of EA in ketamine group was 45% vs15%, respectively. The incidence and severity of EA in the propofol group was group was 20% vs 7.5%, respectively. The incidence and severity of EA in the midazolam group was 15% vs 2.5%, respectively [13]. Compared to our study propofol has a reduction effect on both the incidence and severity of EA. In contrast to our study, they didn’t compare propofol with a placebo.

A study done in Egypt in 2013 on the effectiveness of propofol in the prevention of EA on adenotonsillectomy found that PAEDS at arrival, 5^th^, 15^t^, and 30^th^ minutes was assessed and severity of EA was lower in the propofol group at arrival, 5 and 15 minutes but not at 30 minutes [25]. But the authors did the study only on adenotonsillectomy. However, we included all the surgical procedures and this is what we are expecting.

A study done in Addis Ababa, Ethiopia, in 2021 found that the overall incidence of EA was lower in the propofol group than the non-propofol group (31.1% vs 64.40%), respectively with a p-value of 0.002. The incidence and Severity of EA were also reported at time intervals of 5, 15, and 30 minutes and showed significant differences with a p-value of 0.009, 0.013, and 0.011 [32]. This study was also consistent with the present study in which the severity of EA showed a significant difference between propofol and non-propofol at time intervals of 5^th^, 15^th^. However, in the present study, there was no statistically significant difference between the groups in having EA at 30^th^ minute. The possible reason may be ENT procedures are distressing to the patient and the severity may outlast up to 30 minutes.

### Strength and Limitation of the study

The main strength of this study was, we used the most sensitive and specific scale (PAEDS) for assessing the incidence and severity of EA in which all the study populations were passed under this screen. Besides, the severity of the EA was stratified and assessed at (5^th^, 15^th^ and 30^th^) minute that not studied before. Whereas; small sample size studied is the limitation of study.

## CONCLUSION

We conclude that the administration of prophylactic propofol at the end of surgery in children undergoing surgery under general anesthesia is important for mitigating both the incidence and severity of EA. The reduction of EA could lead to decreased healthcare costs, better patient outcomes, and higher satisfaction rates in patients, parents, and nursing staff.

## Data Availability

All data produced in the present work are contained in the manuscript

## ACKNOWLEDGMENT

We acknowledge data collectors and families/ legal guardians.

## Source of funding

No funding for this work.

## Conflicts of interest

The authors have declared that no competing interests exist.

## ABBREVIATIONS

ASA: American Society of Anesthesiology physical state
EA: Emergence Agitation
GA: General Anesthesia
JUMC: Jimma University Medical Center
OR: Operation Room
PAEDS: Pediatric Anesthesia Emergence Delirium Scale
PACU: Post Anesthesia Care Unit

## Notes

### Competing Interest Statement

The authors have declared no competing interest.

### Clinical Protocols

https://www.researchregistry.com/browse-the-registry#home/

### Funding Statement

This study did not receive any funding

### Author Declarations

Jimma University, institute of health, faculty of medical sciences, institute review board granted ethical approval for this study on September 29, 2022 with protocol number JUIH/IRB/81/22, Jimma, Ethiopia.

### Summary of Updates

great apology for missing one of the coauthor mr. Kanbiro Gedeno Gelebo. now I add him as a coauthor in this preprint. thank you for your consideration.

