## supplementary file for "Effectiveness of Propofol in Mitigation of Emergence Agitation Among Pediatric Patients Undergoing Elective Surgery Under General Anesthesia: Prospective Cohort Study"

**Annexes**

### Annex I: Information Sheet

TITLE OF THE RESEARCH PROJECT: EFFECTIVENESS OF PROPOFOL IN MITIGATION OF EMERGENCE AGITATION IN PEDIATRICS UNDERGOING SURGERY UNDER GENERAL ANESTHESIA IN JIMMA UNIVERSITY MEDICAL CENTER, SOUTHWEST ETHIOPIA, 2022: PROSPECTIVE COHORT STUDY.

**Name of the investigators**:

1. Kuchulo Geremu Gelgelo

2. Orissa Orkissa Tessema
3. Wondu Reta Demissie
4. Gezahegn Tesfaye Mekonin
5. Kanbiro Gedeno Gelebo

This information sheet was prepared with the aim of assessing the effectiveness of Propofol in the mitigation of emergence agitation among pediatrics undergoing elective surgery under general anesthesia.

### Annex-II Procedures and Protocol

To assesses the effectiveness of prophylactic propofol administration at the end of surgery in mitigating the incidence of emergence agitation among pediatrics undergoing elective surgery under GA. First data was collected from all pediatrics that underwent elective surgery under GA and fulfills the inclusion criteria during the study period. Then at the end of each day, we collected all the data and grouped the subjects into propofol and non-propofol using questionnaires.

### Annex-III PAED SCALE

The PAED Scale consists of five characteristics that are each scored by using a 5-point Likert scale.

Make eye contact with a caregiver

Actions are purposeful

Aware of surrounding

Restless

Inconsolable

Two items: The child makes eye contact with the caregiver” and “the child is aware of his or her surroundings,” respectively, are reflections of the child’s state of consciousness and ability to focus attention and to thoughtfully organize external stimuli.

One item, “the child’s actions are purposeful,” addresses changes in cognition that cause the child to behave in a manner outside of what is deemed appropriate. Two additional items, “the child is restless” and “the child is inconsolable,” are measures of psychomotor and emotional behavior, respectively

| The first three items are scored in reverse order | | Items four and five are scored in regular order | |
| --- | --- | --- | --- |
| 4 | Not at all | 0 | Not at all |
| 3 | Just a little | 1 | Just a little |
| 2 | Quite a bit | 2 | Quite a bit |
| 1 | Very much | 3 | Very much |
| 0 | Extremely | 4 | Extremely |

The scores are summed into a final composite score to determine the presence or absence and severity of pediatric emergence delirium. A perfectly calm child scores 0 and extreme agitation corresponds to 20 points. Agitation score: 0-11 no agitation, 12-

15	agitation,	≥16	sever	agitation

### Annex-Iv CONSENT FORM

Study area is Jimma university Medical Center.

Hello! how are you? I am **-----------------------------**here to collect information on assessing the effect of prophylactic administration of propofol on incidence and severity of emergence agitation in pediatric patients having elective surgery under general anesthesia at Jimma university medical center from August 5/2022-October 30/2022. A study will be conducted by Kuchulo Geremu Gelgelo, Orissa Orkissa Tessema, Wondu Reta Demissie and Gezahegn Tesfaye Mekonnen that are lecturers in department of anesthesia.

The study is aimed at improving health planning and safe emergence during recovery from general anesthesia since the study is not linked with any financial aid there are no direct incentives paid as a result of your child taking part in the study. I would like to assure you, that your child's name will not be written on this form and all the information gathered will be kept strictly confidential. Are you willing to participate in the study?

1/ Yes, Sign___________	2/No

Thank you for your participation!

For more information and question contact:

1. Kuchulo Geremu Gelgelo

2. Orissa Orkissa

3. Wondu Reta Demissie

4. Gezahegn Tesfaye Mekonin

5. Kanbiro Gedeno Gelebo

### ANNEX-V: QUESTIONNAIRE

Questionnaires were developed for the collection of data on the study of “assessing the effectiveness of prophylactic administration of propofol in mitigation of emergence agitation among pediatric patients between 2-14 years old and had elective surgery under general anesthesia in Jimma university medical center in Jimma, Ethiopia.” **Instruction:** For each question, please encircle the possible response/s/

Patient code:

Part one: Socio demographics of the child:

| Serial number | Question | Response | |
| --- | --- | --- | --- |
| 101 | Age |  | |
| 102 | Sex | Male | Female |
| 103 | Weight (kg) |  | |

Part two: Preoperative parameters

| 201 | ASA statues | ASA I | ASA II | | |
| --- | --- | --- | --- | --- | --- |
| 202 | Write if any  Coexisting disease | _________________________ | | | |
| 203 | Diagnose | __________________________ | | | |
| 204 | Type of surgery | ENT  Ophthalmic  Abdominal  Urology | | | Orthopedics  Neuro  Thoracic  Specify if other----------- |
| 205 | Premedication | YES | |  | NO |
|  | If yes to the above question what medication is given? | Atropine dexamethasone  Ketamine | | Diazepam  Midazolam  Specify if other ------------- | |

Part three: Intraoperative parameters

| 302 | Type of induction agent | Propofol  Ketamine | Ketofol  Inhalation agent |
| --- | --- | --- | --- |
| 303 | Analgesics used | Paracetamol  Fentanyl  Pethidine | Morphine  Tramadol  Diclofenac  Other specify |
| 304 | Maintenance of anesthesia | Halothane  Isoflurane  Sevoflurane | propofol infusion  Other agents ---- |
| 305 | Prophylactic propofol give at the end of surgery | Yes  No |  |
|  | If yes to the above question write the dose |  |  |
| 306 | Duration of surgery |  |  |
| 307 | Duration of anesthesia |  |  |

Part four: Emergence characteristics

402. Measures of presence and severity of emergence agitation as PAED scale. At: 05, 15, and 30 minutes.

| Behavior | Not at all | just a little | quite a bit | very much | Extremely | at 05 minute | at 15 minute | at 30 minute |
| --- | --- | --- | --- | --- | --- | --- | --- | --- |
| Make eye contact with the caregiver | 4 | 3 | 2 | 1 | 0 |  |  |  |
| action is purposeful | 4 | 3 | 2 | 1 | 0 |  |  |  |
| aware of surrounding | 4 | 3 | 2 | 1 | 0 |  |  |  |
| Restless | 0 | 1 | 2 | 3 | 4 |  |  |  |
| Inconsolable | 0 | 1 | 2 | 3 | 4 |  |  |  |
| Total score |  |  |  |  |  |  |  |  |
